# Systematic policy and evidence review to consider how dementia education and training is best delivered in the social care workforce, and how policy does or can enable its implementation in England

**DOI:** 10.1101/2024.08.24.24312532

**Authors:** Saskia Delray, Sube Banerjee, Sedigheh Zabihi, Madeline Walpert, Karen Harrison-Dening, Charlotte Kenten, Yvonne Birks, Clarissa Giebel, Mohammed Akhlak Rauf, Sally Reynolds, Claudia Cooper DeNPRU-Queen Mary

## Abstract

**Background:** Very many social care clients have dementia, but few social care workers receive dementia-specific training.

**Objective:** To systematically review dementia training interventions for social care, review past policies and hold stakeholder workshops considering how future policy can support quality dementia training in social care.

**Methods:** We searched electronic databases, November 2015 to February 2024, including studies describing dementia training and support interventions for social care workers, assessing risk of bias with the Mixed Methods Appraisal Tool. We reviewed English policies January 2015 to April 2024 to identify social and policy contexts relevant to dementia training. We consulted home care and care home stakeholders regarding how findings could inform future policy.

**Results:** We included 56 studies (50 in care homes, 6 in home care). There was good quality evidence that dementia training interventions in care homes that engaged staff “champions” to integrate practice-based learning reduced agitation, neuropsychiatric symptoms and antipsychotic prescribing and improved life quality of residents with dementia. One study found this approach was cost-effective. In home care, evidence was limited; group training was valued, and improved staff sense of dementia care competence in one study. We identified 27 policies and related documents; and consulted 18 stakeholders. Stakeholders supported mandatory dementia training but considered implementation very challenging in current economic contexts.

**Conclusions:** We found strong evidence for dementia training in care homes, but a relative lack of research in home care. Policy options identified to implement evidence require investment, which could deliver substantial savings across health and social care.

## Introduction

Around 982,000 people in the UK live with dementia, often undiagnosed. Many require care which employs specific skills and knowledge (1). A million people work as care workers in UK social care (2). Nearly half of home care residents have significant cognitive impairment, a third have a dementia diagnosis (3) and over three-quarters of care home residents have dementia (4). Care workers are low-paid, often undervalued and expected to provide skilled care to people with dementia with little or no training (5–7). Dementia care should be personalised, enabling, inclusive and collaborative (8,9), but current provision is inconsistent in quality (5).

We aimed to update a 2015 review of effectiveness of social care sector dementia training, which found most effective programmes comprised 12+ hours of training by experienced facilitators; were tailored to staff needs, included active participation, and underpinned practice-based learning with theory (10). This update is timely. The 2015 *Dementia Training Standards Framework* benchmarked training for the health and care workforce (11); benchmarks which contemporary provision mostly failed to meet (12). Developing an appropriately skilled social care workforce is a policy priority (13). The previous UK government planned to build upon the Care Certificate with a careers pathway and new qualifications (14,15). In July 2024, the new government announced long-term plans for a National Care Service underpinned by national standards, to deliver consistency of care (16). To inform future policy regarding dementia training for social care, we aimed to review policy and academic evidence, adapting a previous framework (17) to answer the following research questions:

1. What is the social and policy context determining how dementia training is delivered to social care workers in England?
2. What is the current evidence on how dementia training is best delivered to improve care quality and care worker wellbeing?
3. How do stakeholders consider current evidence might inform future policies to positively change social care workforce training?

## Methods

### 1. Defining social and policy context

This review was conducted within the NIHR Dementia and Neurodegeneration Policy Research Unit - at Queen Mary (DeNPRU-QM), overseen by a study group comprising: a psychiatrist, two social care academics, two lived experience members, two researchers and two senior nurses. We consulted the wider DeNPRU-QM group, and an external oversight group with policy and clinical expertise in November 2023.

The study group agreed sources and terms to search policy and grey literature (from 2015) to respond to RQ1. SD and CC conducted the search (Supplementary Table 1a) using terms: “social” and “training” and “dementia” to identify potentially relevant documents. SD and CC independently identified documents in April 2024, then discussed those to include, based on relevance to the research question, consulting the study group iteratively. They tabulated findings, meeting regularly with the study group to discuss emerging data, and develop descriptions of contexts and problems as our empirical and conceptual understanding evolved during the review (17).

### 2. Systematic review of academic evidence

We prospectively registered the review protocol [PROSPERO: CDR42024509026].

#### Search strategy and selection criteria

We reviewed the following electronic databases from 01.12.15 (updating (10)) to 20.02.2024: Pubmed, Embase, Scopus, CINAHL and the Cochrane library. We planned the search strategy around RQ2. Terms related to ‘education’/‘training’, ‘staff’, and ‘dementia’ were combined with the Boolean operators ‘AND’ and ‘OR’, linking search terms within concepts. Supplementary Table 1b outlines the Pubmed search; we developed similar strategies for other databases. We asked experts to identify unpublished studies.

We included primary research (quantitative and/or qualitative) studies, in English, reporting the effectiveness of training/education interventions aiming to develop dementia-specific knowledge, values and skills to improve dementia care, delivered to social care workers in care home or home care settings. We excluded individual case studies, dissertations and meeting abstracts. Because our focus was on training to manage dementia symptoms, we excluded studies evaluating training limited to care for co-morbid conditions, or end-of-life care in the context of dementia rather than dementia symptom management.

#### Data extraction

Using Covidence software, two reviewers independently assessed potential studies against inclusion criteria, in an initial title and abstract screening, then full paper review, discussing divergences. Tables 1-2 show the data extracted. We used Kirkpatrick’s model to classify outcomes: learner’s reaction to/ satisfaction with training (Level 1); knowledge, skills, confidence, and attitudes (Level 2); staff behavior or practices (Level 3); client/staff wellbeing (Level 4)(18).

**Table 1:**
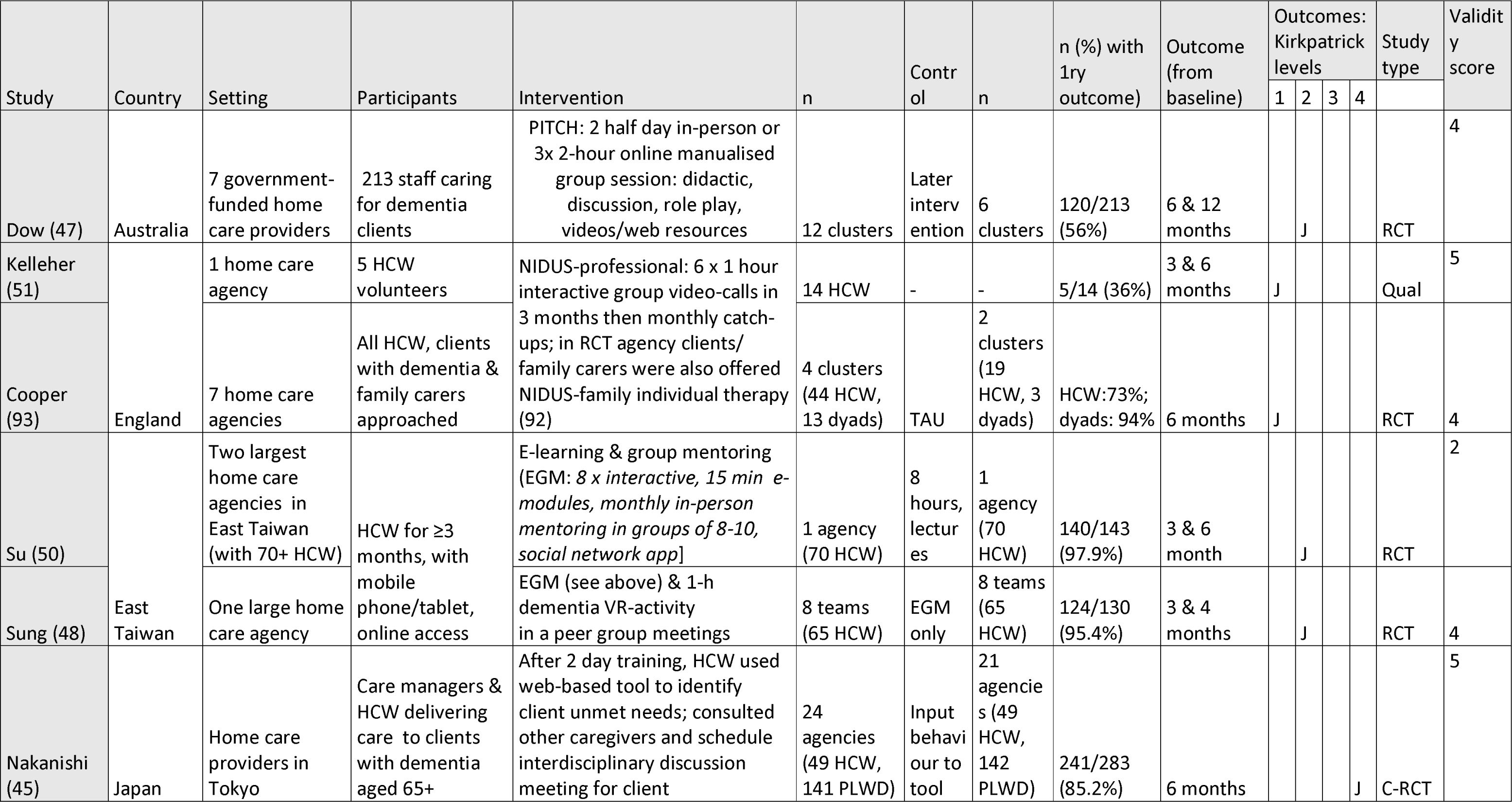
Characteristics of included studies based in home care.

| Study | Country | Setting | Participants | Intervention | n | Contr<br>ol | n | n (%) with<br>1ry<br>outcome) | Outcome<br>(from<br>baseline) | Outcomes:<br>Kirkpatrick<br>levels |  |  |  | Study<br>type | Validit<br>y<br>score |
| --- | --- | --- | --- | --- | --- | --- | --- | --- | --- | --- | --- | --- | --- | --- | --- |
|  |  |  |  |  |  |  |  |  |  | 1 | 2 | 3 | 4 |  |  |
| Dow (47) | Australia | 7 government-funded home care providers | 213 staff caring for dementia clients | PITCH: 2 half day in-person or 3x 2-hour online manualised group session: didactic, discussion, role play, videos/web resources | 12 clusters | Later intervention | 6 clusters | 120/213 (56%) | 6 & 12 months |  | J |  |  | RCT | 4 |
| Kelleher (51) | England | 1 home care agency | 5 HCW volunteers | NIDUS-professional: 6 x 1 hour interactive group video-calls in 3 months then monthly catch-ups; in RCT agency clients/ family carers were also offered NIDUS-family individual therapy (92) | 14 HCW | - | - | 5/14 (36%) | 3 & 6 months | J |  |  |  | Qual | 5 |
| Cooper (93) |  | 7 home care agencies | All HCW, clients with dementia & family carers approached |  | 4 clusters (44 HCW, 13 dyads) | TAU | 2 clusters (19 HCW, 3 dyads) | HCW:73%; dyads: 94% | 6 months | J |  |  |  | RCT | 4 |
| Su (50) | East Taiwan | Two largest home care agencies in East Taiwan (with 70+ HCW) | HCW for ≥3 months, with mobile phone/tablet, online access | E-learning & group mentoring (EGM: 8 x interactive, 15 min e-modules, monthly in-person mentoring in groups of 8-10, social network app] | 1 agency (70 HCW) | 8 hours, lectures | 1 agency (70 HCW) | 140/143 (97.9%) | 3 & 6 month |  | J |  |  | RCT | 2 |
| Sung (48) |  | One large home care agency |  | EGM (see above) & 1-h dementia VR-activity in a peer group meetings | 8 teams (65 HCW) | EGM only | 8 teams (65 HCW) | 124/130 (95.4%) | 3 & 4 months |  | J |  |  | RCT | 4 |
| Nakanishi (45) | Japan | Home care providers in Tokyo | Care managers & HCW delivering care to clients with dementia aged 65+ | After 2 day training, HCW used web-based tool to identify client unmet needs; consulted other caregivers and schedule interdisciplinary discussion meeting for client | 24 agencies (49 HCW, 141 PLWD) | Input behaviour to tool | 21 agencies (49 HCW, 142 PLWD) | 241/283 (85.2%) | 6 months |  |  |  | J | C-RCT | 5 |

**Table 2:**
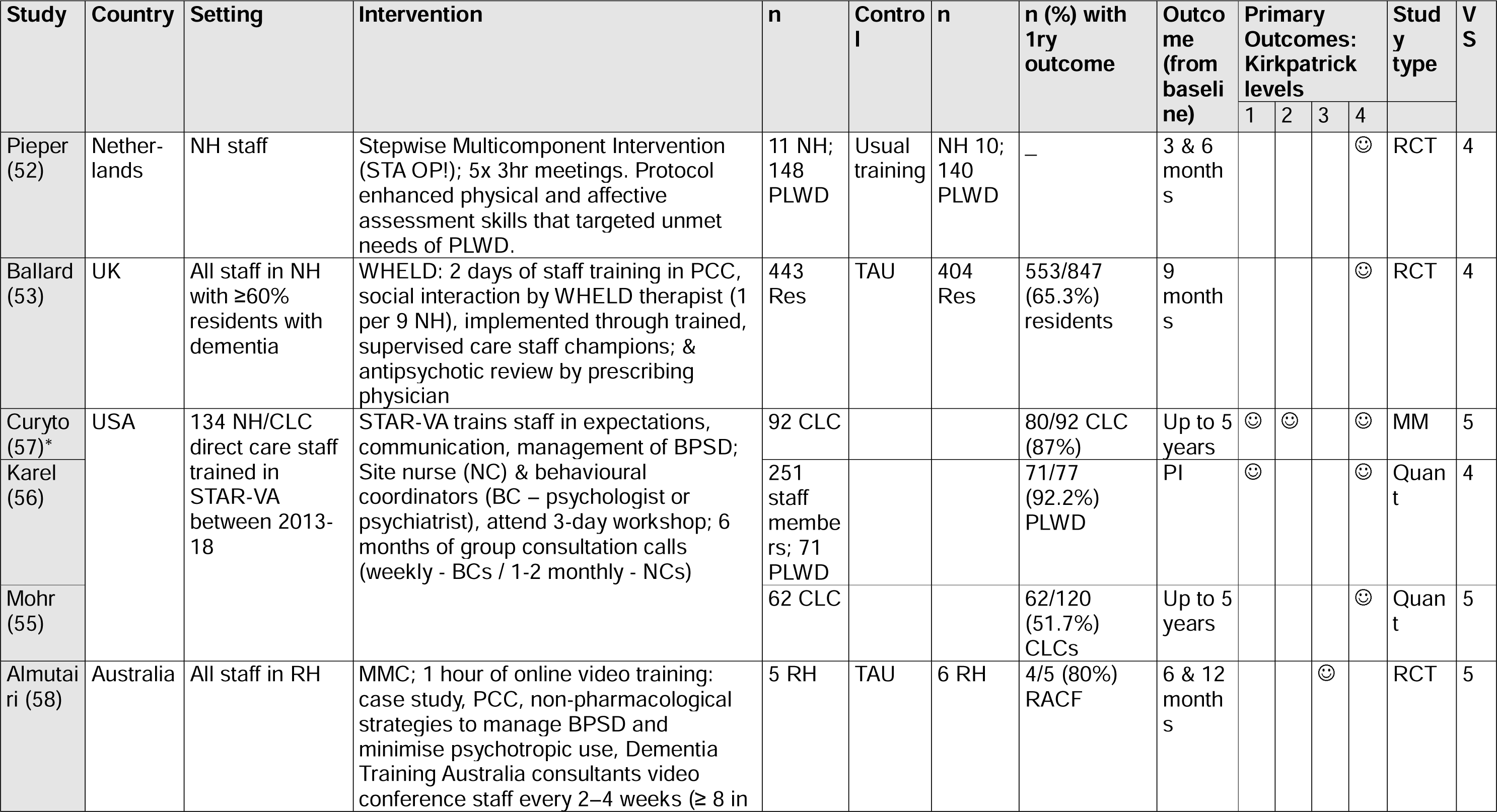
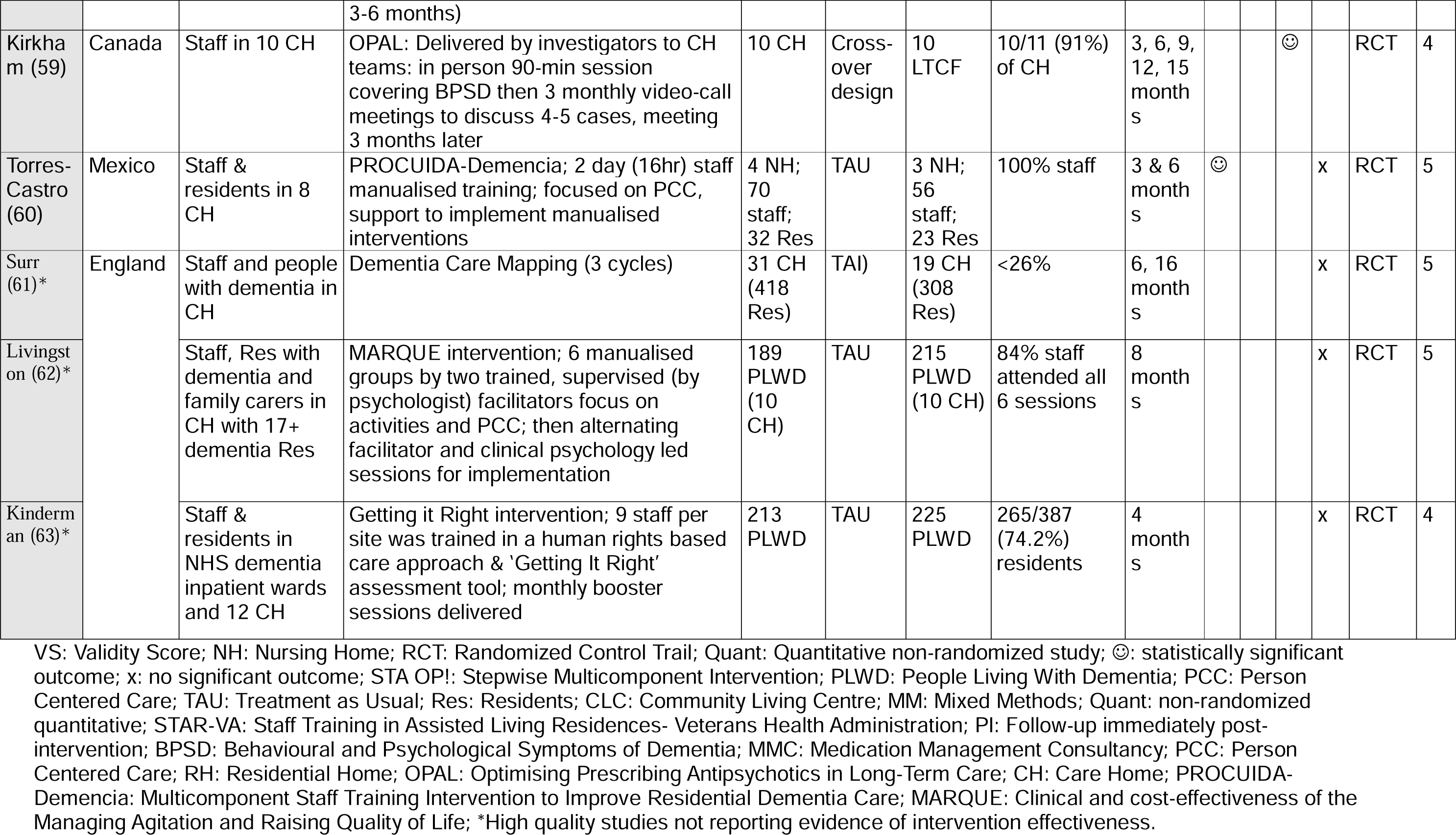
Characteristics of included studies based on care homes – studies meeting criteria for higher priority.

#### Analysis

SD and CC independently assessed risk of bias of included studies using the Mixed Methods Appraisal Tool (MMAT)(19). We calculated rater percentage agreement, then resolved discrepancies through discussion. We prioritised evidence from studies scoring 4+ on MMAT (indicating high quality), that were Randomised Controlled Trials (RCTs) (or implementation studies of interventions for which RCT evidence is published), which, in line with our aim to identify evidence that may inform future training programmes, reported a significant finding on a main outcome in a between-group comparison. We descriptively synthesised findings for care home and home care settings by Kirkpatrick’s levels (18).

### 3. Stakeholder consultation

To answer RQ3, we held two workshops in April and June 2024 with (1) home care managers and workers and (2) care home workers and managers. We presented groups with findings in accessible formats (Supplementary Tables 5-6) and asked them to consider how evidence-supported interventions might be used to lever change to improve care quality and potential outcomes of any proposed policy changes. SD/CK synthesised the discussion and fed this back to attendees, inviting corrections or further comments.

## Results

### 1. Defining social and policy context

We identified 27 relevant policy and related documents (Supplementary Table 8), from which our study group summarised key policies and contexts and problems to explore in stakeholder groups (RQ3) (Figure 1). Identified documents advocated for quality training as an essential part of providing good quality (20), culturally competent (21), personalised dementia care (22) (23). The Dementia Core Skills Education and Training Framework delineates learning objectives for care workers; with 3 Tiers, Tier 2 the level expected of care workers and Tier 3 expected of managers and leaders (24). NICE dementia guidelines recommend in-person, case-based training encompassing strategies to reduce distress and antipsychotic use, promote freedom of movement, and have difficult conversations (25). Training expectations that new social care staff are expected to work towards completing in their first three months are operationalised through the Care Certificate (26), which two-thirds of care workers in England engage with; a qualification based on it (27) with funding for 37,000 care workers to enrol was developed by the previous government (28). Dementia training is not mandatory for social care workers (29), but the All-Party Parliamentary Group (APPG) for Dementia has called for Tier 2 training for care workers to be mandatory (30). A new Care Workforce Pathway (31), advocates leadership training for managers (32) (33). Training needs have been cited around digitisation (of care records) (34)(35) and wider digital skills (36). Health and social care integration implementation work has proposed greater cross-sector training, including support for extended roles in skills traditionally performed within healthcare (13). An integrated skills passport has been proposed across health and social care (15).

**Figure 1:**
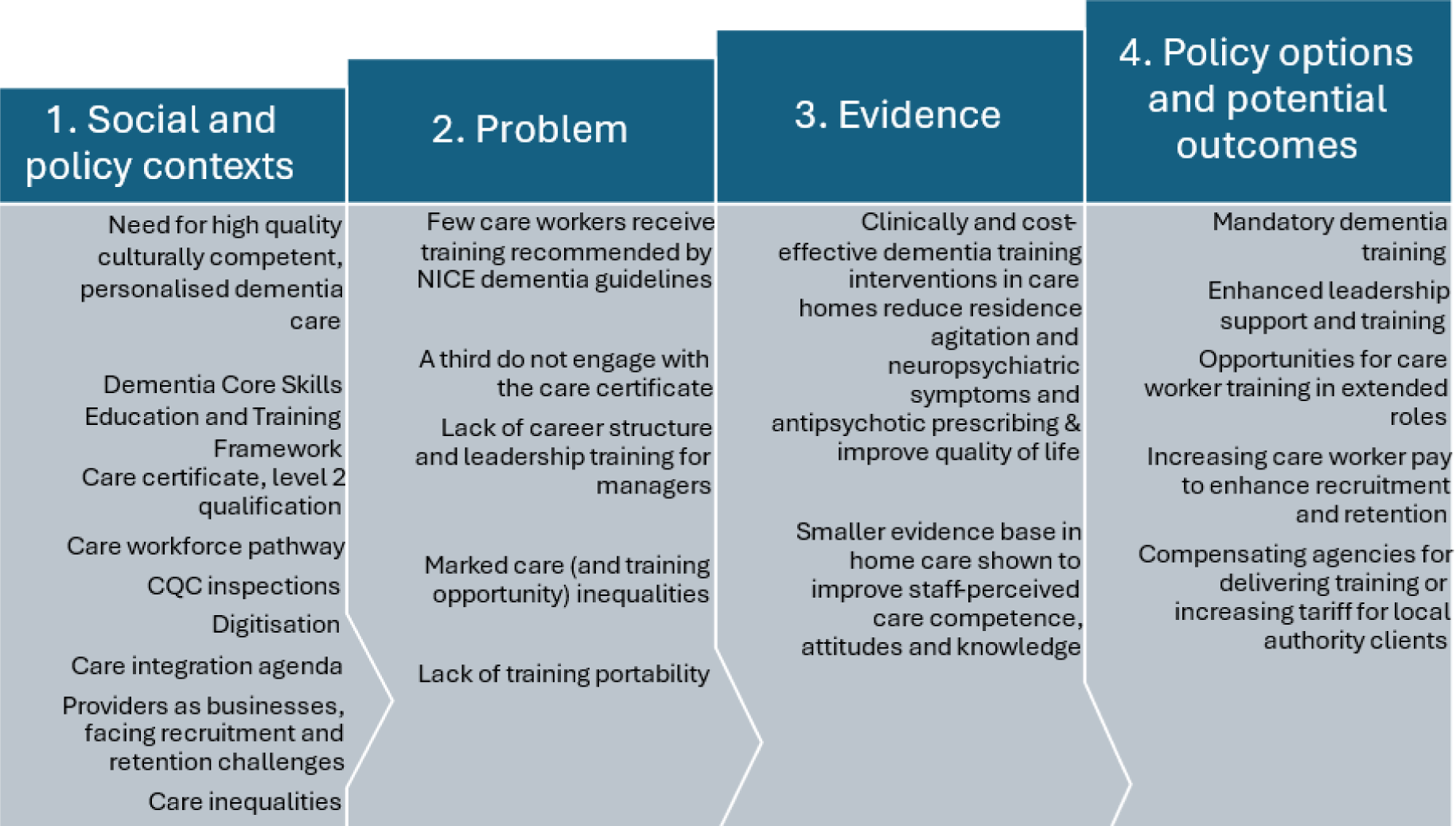
Summary of evidence developed across stages of our review

Care Quality Commission (CQC) inspections can drive quality improvement (37). Providers are private businesses facing recruitment (including international) and retention challenges (38) (39) and adverse economic conditions (40), but training could improve retention (41), care quality and help meet needs of informal carers for support (42). There are geographic and socioeconomic inequalities in resources with lower CQC ratings linked to fewer resources (likely including training) potentially due to fewer self-funded clients in care homes (43) (44).

### 2. Systematic review

#### Search results (see PRISMA, Figure 2)

We identified 5,939 studies in our electronic search and after screening 56 studies (6 in home care settings, 50 in care homes) met our inclusion criteria.

**Figure 2:**
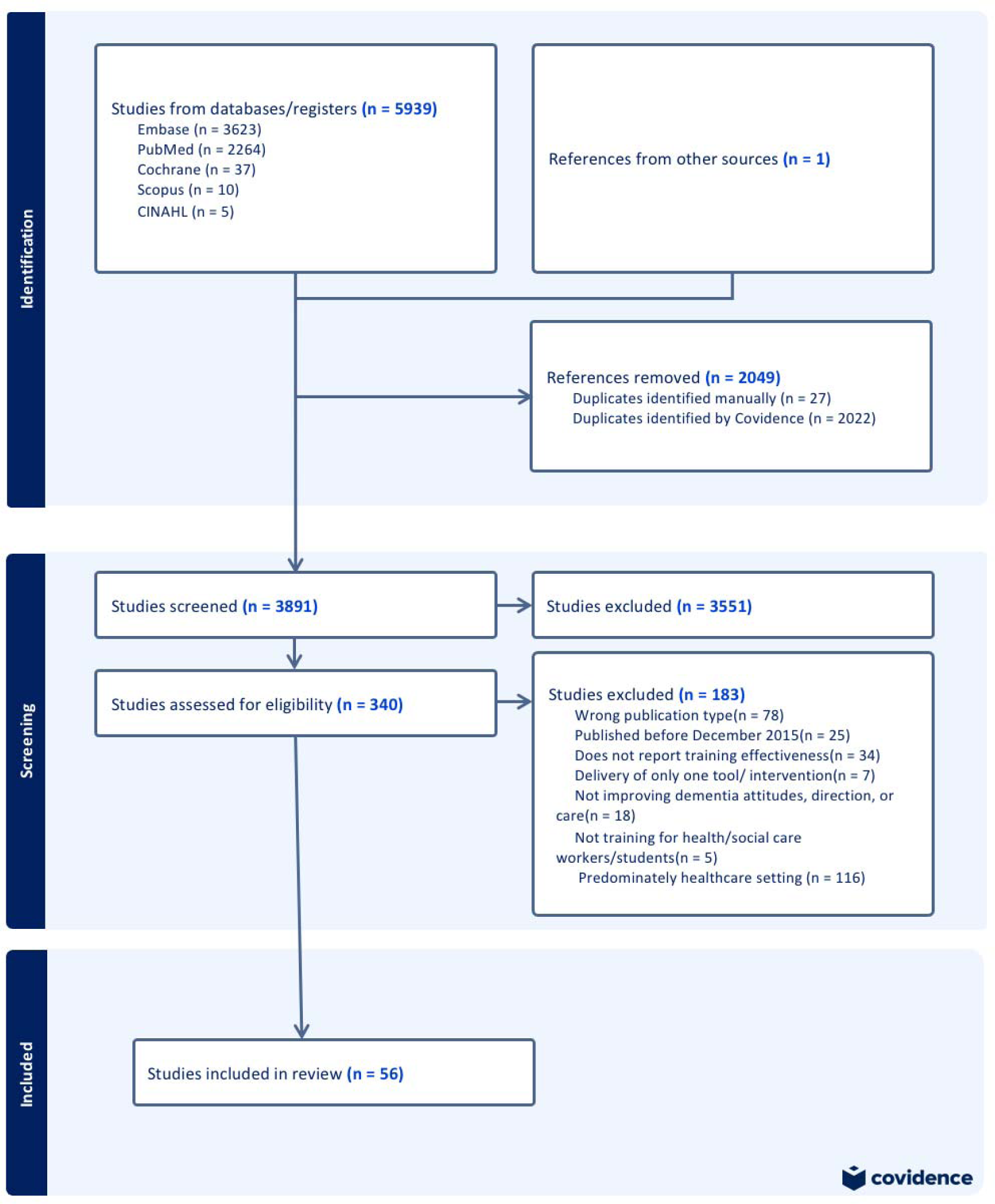
PRISMA (Preferred Reporting Items for Systematic Reviews and Meta-Analyses) diagram of included and excluded studies.

#### Description of studies

22/56 (39%) studies were rated 5/5 on the MMAT, the lowest risk of bias; thirteen were rated 4. Rater agreement was 90.7% (245/270) for MMAT items. Supplementary Table 2 shows MMAT ratings. Thirteen studies met our criteria for priority evidence (see methods for criteria).

#### Home Care Studies (n=6; Table 1)

Studies were conducted in Taiwan (n=2), Australia (n=1), England (n=2) and Japan (n=1). 4/6 studies rated as priority evidence, described four interventions.

**Level 4 evidence:** In the Japanese Behaviour Analytics & Support Enhancement (BASE) program (45), care managers (social workers) and home care workers attended a two day training event with expert trainers. Home care workers used a web tool to identify client unmet needs and record behaviours that challenged; they sought other care workers’ views and organised interdisciplinary case conferences for included clients. The control group continued standard client support, including monthly meetings with care managers mandated by the Japanese Public Long Term Care Insurance Plan. In the primary analysis, challenging behaviour (Neuropsychiatric Inventory—Nursing Home version (NPI-NH)) reduced in the intervention (mean score: 18.3 to 11.2) relative to the control group at six months (11.6 to 10.8; P<0.05).

**Level 2 evidence:** The Promoting Independence Through quality dementia Care at Home (PITCH) group training intervention delivered by an expert trainer, focused on valuing home care workers, and person-centred care skills (46). Delivery was in-person or, during the COVID-19 pandemic, online (47). In a stepped wedge RCT, the primary outcome, home care worker Sense of Competence in Dementia Scale (SCIDS) (F=4.48, p=0.04), and Dementia Attitudes Scale (DAS) and Dementia Knowledge Assessment Scales (DKAS) improved significantly in the intervention relative to control group.

In the E-learning and Group Mentoring with Virtual Reality (EGM-VR) study, Sung (48) compared EGM with and without an hour of VR training. EGM involved eight, 15-minute interactive e-modules, monthly in-person mentoring in groups of 8-10, and social network app support. The VR condition immersed participants in perspectives of people living with dementia, with a facilitated 50-minute discussion. Over three months, DKAS (P<0.001), Approaches to Dementia Questionnaire (ADQ (P<0.001), the Jefferson Scale of Empathy (P=0.001) and SCIDS (p<0.001) scores increased in the EGM + VR relative to EGM only group.

**Level 1 evidence:** A UK, RCT feasibility study evaluated six, interactive, manualised, one-hour, fortnightly group video-calls focused on valuing and supporting staff, developing communication skills, empathy with clients and strategies for managing behaviours that challenge, over three months with monthly catch-up group calls to discuss implementation. Intervention development drew on the PITCH intervention (47). The primary outcome was intervention feasibility and acceptability, with 65.9% of sessions attended. The average cost per agency of training provided was £1,606 (49).

**Evidence from studies rated as lower priority:** Su (50) evaluated EGM alone, relative to usual care. DKAS, ADQ, SCIDS scores significantly improved in the intervention compared to control group. Kelleher published a qualitative pilot study, which like the feasibility RCT of the same intervention above, was found to be acceptable (51).

#### Summary of Home Care evidence

- We found good quality evidence from single studies that two days of group, expert training, client behaviour monitoring and monthly case conferences were better than case conferences alone at reducing neuropsychiatric symptoms; six hours of group training with an expert trainer improved staff sense of competence, attitude, knowledge and empathy in dementia care.

### Care home Studies

#### Higher priority Evidence (n=8/50 Table 2)

Studies were conducted in the Netherlands (n=1), UK (n=1), USA (n=3), Australia (n=1), Canada (n=1) and Mexico (n=1) and evaluated six interventions.

**Level 4 evidence:** A Dutch study compared usual training to a Stepwise Multicomponent Intervention (STA OP!) involving five, 3-hour meetings teaching step-wise management of behavioural symptoms of distress in people with severe dementia (52). This considered basic care needs, then pain and physical needs, affective needs, non-pharmacological comfort interventions, trials of analgesia and finally antipsychotic medication. Key messages were reiterated in team meetings, and a coordinator visited homes weekly and evaluated fidelity. This cluster RCT reported a significant reduction in agitation, the primary outcome in the intervention versus control (Cohen-Mansfield Agitation Inventory (CMAI) mean difference −4.07, 95% CI= −7.90 to −0.24, P=0.02), NPI-NH and depression scores. Though the intervention was intended to support all residents, in practice only a small proportion, likely those with most challenging behaviours were discussed.

The UK WHELD intervention described a 9-month in-person training course in person-centred care, social interaction and antipsychotic medication reviews (53). An Occupational Therapist (1 per 9 nursing homes) visited staff and residents for 2 days and identified WHELD champions who received one day of training per month, to cascade training to staff, and guide implementation. The cluster RCT showed significant improvement in quality-of-life in the treatment versus control arm (DEMQOL-Proxy mean difference 2.54, 95% CI 0.81, 4.28). WHELD cost £8,627 more per nursing home to deliver compared to usual training, but client health and social care costs were lower in the intervention than usual training group. There were statistically significant benefits in agitation (CMAI) and NPI-NH, and positive care interactions. Intervention adherence was not reported.

Three studies evaluated the US Staff Training in Assisted Living Residences (STAR) intervention, which reduced resident distress and challenging behaviours and increased staff job satisfaction over eight weeks in an earlier RCT (54). STAR involves a 3-day staff workshop in expectations, communication and managing distress behaviours in people with dementia, attended by staff champions who then cascade training to sites, supported through ongoing training to implement the approach. In implementation studies, weekly consultation video calls with the STAR team supported staff to apply learning. Mohr (55) found that over 6 months, STAR was associated with lower staff injury rates (b=-0.04, Standard Error (SE)=0.02, p=0.04). Curyto (55) and Karel (56) reported that over six years, STAR reduced distressed behaviour (DBID) (t test (t)= 12.9, P<0.001, Cohen’s d= 0.8), depression (CSDD) (t=14.0, P<0.001, d=0.8), anxiety (RAID) (t=13.4, P<0.001, d= 0.6) and CMAI-short form (CMAI_SF) (t=14.7, P<0.001, d= 0.9) scores. There was significant improvement in self-perceived staff confidence after the STAR-VA training and consultation compared to usual training (57).

**Level 3 evidence:** Two studies tested interventions that sought to decrease antipsychotic prescribing. A six-month Australian program involved one hour of e-learning, then eight video conferences with a focus on medication audits and non-pharmacological strategies, delivered by Dementia Training Australia consultants to care home staff. In a cluster RCT, antipsychotic use was lower relative to the control group at six months (IRR, 0.56, 95% CI, 0.32-0.99; P=0.048), but not at 12 months (58). A five-month Canadian program, comprising a 90-minute in-person group staff training session with three subsequent, monthly interdisciplinary video calls, case-based discussions, to reduce inappropriate prescribing of antipsychotics in long-term care. There was a 16.1% relative reduction in the inappropriate antipsychotic prescribing rate (receiving antipsychotics without a diagnosis of psychosis) over 12 months (SD=2.8%, P< 0.0001) (59).

**Level 1 evidence:** A Mexican feasibility RCT evaluated two days of on-site group staff training around understanding dementia, teamwork, and using psychosocial interventions with support to put learning into practice over 12 weeks. Medical staff attending care homes received training in antipsychotic reviews. Fidelity, acceptability and satisfaction scores were high (60).

##### Summary of high priority evidence

- Three studies provided good quality evidence that training in person-centred care approaches reduced resident agitation and neuropsychiatric symptoms and improved quality of life. Methods to embed learning in team meetings and care included training and supporting staff champions, video-call case conferences with project experts, and project coordinator visits. In one study evaluating cost-effectiveness, WHELD cost £8,627 more than usual training; but was cost-effective from a health and social services perspective. Two studies provided good quality evidence that interventions comprising initial training, then video-conference case-based discussions reduced antipsychotic prescribing.

### Lower priority Evidence

#### Low risk of bias RCTs reporting negative findings (n=3, Table 2)

Surr (61) evaluated Dementia Care Mapping (DCM) training and three DCM cycles, the first supported by an expert. Livingston (62) evaluated six manualised interactive group sessions by a supervised graduate facilitator, then monthly supervision, alternating between a clinical psychologist and graduate facilitator. Neither study reported significantly improved clinical (agitation) or cost-effectiveness. Kinderman (63) evaluated a one-day staff training offer framed in a human-rights based tool, then three-monthly booster sessions and found no between-group difference in quality of life.

#### Higher risk of bias-rated RCTs (n=9) (Supplementary Table 3)

Two interventions involving up to 12 hours of expert training and group, case-based, discussion, reduced behavioural distress (64,65) and psychotropic prescribing (64). The CHAnging Talk staff education program (CHAT) (66,67) and CHAT Online (CHATO) (68) comprised of three, one-hour, group/online interactive training modules; they reduced ‘elderspeak’ and antipsychotic use compared to state averages (59); CHATO also improved staff communication (68).

Digital WHELD (eWHELD), involved a two-month virtual coaching phase plus five 25-minute e-modules focusing on person-centred care. McDermid (69) compared eWHELD with and without virtual coaching. Resident wellbeing, positive activities, staff attitudes, staff hope and personhood, improved in the group receiving virtual coaching. Testad (70) evaluated a seven month intervention by research nurses, which did not reduce restraint use. A staff training for people with advanced dementia did not reduce unplanned hospital transfers, though low staff participation was reported. (71).

#### Non-RCT evidence (n=30) (Supplementary Tables 4-5)

We rated 19 non-RCT studies as high quality (Supplemental Table 3) and 11 as lower quality (Supplemental Table 4). We summarise here studies rated as low risk of bias, reporting effectiveness.

**Level 4:** In-person interventions, involving group workshops focused on person-centred care, were associated with improvements in neuropsychiatric symptoms (72), agitation (73), fewer clinical symptoms (74), and with resident quality of life and reduced carer burden in a Canadian training program involving development of individualised communication plans (75). A German study found that a “train the trainer” communication intervention reduced resident depressive symptoms (76).

**Level 3:** Hanson’s findings echoed those from RCT evidence, that with training, inappropriate antipsychotic prescribing reduced; cost per prescription avoided was $5,679 (77). Project ECO-AGE, was a US intervention involving online video case discussions with clinical experts bi-weekly, over 18 months. Residents in ECO-AGE facilities were 75% less likely to be physically restrained and 17% less likely to be prescribed antipsychotic medication compared to control facilities (78).

**Level 2:** A Chinese programme, delivered face-to-face, weekly for eight weeks was associated with improvement in sense of competence in dementia care (79,80). Further studies showed significant improvements in staff knowledge following in-person training (79)(81)(82)(83)(84)(85).

**Level 1:** Four studies reported good acceptability of interventions (86,87) (88) (89).

### 3. Stakeholder consultation

We consulted 18 care managers and workers; meeting on 30.4.24 in-person with nine home care managers and workers from two franchises; and 25.6.24, online, with eight care home managers and workers from four organisations. Both groups raised the policy option of mandatory dementia training and concerns about implementing this without additional funding, referencing resource challenges delivering Oliver McGowan Mandatory training on learning disability and autism (90). Care workers pay and agency reimbursement for local authority clients were perceived as major barriers to implementing any training, though staff valued training opportunities. The home care group discussed how training could enhance retention and job satisfaction.

When asked how evidence-based interventions might work in practice (Supplementary Tables 5-6), home care staff gave workforce geographical dispersal and high turnover as reasons for preferring online, regular training, with most considering NIDUS-professional the most practical option to implement of the options presented (49). Care home staff favoured approaches used in interventions, such as WHELD (53), of training staff “champions” to cascade training, so content could be tailored to the home’s culture.

Figure 1 summarises policy options discussed.

## Discussion

Increasing dementia training for the (circa) million social care workers caring for people living with dementia is critical. Policies have outlined frameworks for learning objectives, qualifications and regulatory systems that could drive this. There is a good evidence base for dementia training in care homes, but significant evidence gaps in home care. This sector presents a different care delivery context; lone working, domiciliary settings and working alongside family carers require specific expertise. We found good quality evidence that training and supporting care home staff “champions” to integrate practice-based learning reduced agitation, neuropsychiatric symptoms and antipsychotic prescribing and improved life quality of residents with dementia. In home care, group training was valued, and improved staff sense of dementia care competence in one study (47).

In July 2024, Skills for Care (91) called for mandatory dementia training. Stakeholders we consulted supported such calls but considered implementation unfeasible in current economic and workforce contexts. Policy options that the new government might consider to implement this training base would require investment, but could deliver substantial savings across health and social care in care homes (53). There was no cost-effectiveness data published for home care training, though one intervention that staff valued cost £1,606 per agency, suggesting that if relatively modest health and social care savings were accrued, training could be cost-effective.

Though challenging, widespread implementation of dementia training across social care is necessary to equip professionals to deliver the high quality, personalised care that people with dementia and their family carers need. Evidence to drive this is available for care homes but would most likely require regulatory enforcement and investment if care home residents with dementia are to benefit. Half a million people living in UK care homes, and the majority have dementia so successful implementation would support many. In the home care sector, definitive intervention trials are needed.

## Data Availability

All data produced in the present study are available upon reasonable request to the authors

