## Supplemental material for "Systematic policy and evidence review to consider how dementia education and training is best delivered in the social care workforce, and how policy does or can enable its implementation in England" (from another version of this preprint: version 2, 2024-08-26)

**Supplementary Materials**

**Supplementary Table 1a: Sources search for policy and grey literature**

|  | Organisation websites searched |
| --- | --- |
| Governmental and arms length bodies | Department of Health and Social Care (DHSC)  Health Education England  Skills for Care  National Institute for Health and Care Excellence (NICE)  All-Party Parliamentary Group (APPG) on Dementia  NHS England  Care Quality Commission (CQC) |
| Third sector and industry | Alzheimer’s Society  Homecare association  Care England  Directors of Adult Social Services  Unison  Care Provider Alliance |

**Supplementary Table 1b: Search strategy for Pubmed (dates 2015- filter applied)**

| Terms |  |
| --- | --- |
| Education | (educat*[Title/Abstract] OR training[Title/Abstract] OR staff development[Title/Abstract] OR professional development[Title/Abstract] OR CPD[Title/Abstract] OR skills training[Title/Abstract] OR curricul*[Title/Abstract] OR learn*[Title/Abstract] OR teach*[Title/Abstract] OR workshop[Title/Abstract] OR module[Title/Abstract]) |
| Staff | ((professional[Title/Abstract] OR staff[Title/Abstract] OR worker[Title/Abstract] OR workforce[Title/Abstract] OR paid carer[Title/Abstract] OR aide[Title/Abstract] OR care worker[Title/Abstract] OR physician[Title/Abstract] OR doctor[Title/Abstract] OR student[Title/Abstract] OR nurse[Title/Abstract] OR therapist[Title/Abstract] OR social worker[Title/Abstract]) |
| Dementia | (dementia[Title/Abstract] OR "Frontotemporal dementia"[Title/Abstract] OR korsakof*[Title/Abstract] OR binswanger[Title/Abstract] OR "Progressive Supranuclear palsy"[Title/Abstract] OR alzheimer*[Title/Abstract] OR dement*[Title/Abstract] OR "Korsakoff Syndrome"[Title/Abstract] OR "Wernicke's Encephalopathy"[Title/Abstract] OR "Huntington's Disease"[Title/Abstract] OR "Multi Infarct"[Title/Abstract] OR "lewy bodies"[Title/Abstract] OR "Lewy Body Disease"[Title/Abstract] OR "kluver-bucy syndrome"[Title/Abstract] OR "Vascular dementia"[Title/Abstract] OR "Creutzfeldt-Jakob Syndrome"[Title/Abstract] OR "Alzheimer's Disease"[Title/Abstract])) |

***Supplementary Table 2: Quality evaluation of included studies, rated using the Mixed Methods Appraisal Tool (MMAT)***

|  | 1. **Qualitative** | | | | | | | | 2. ***Quantitative RCT*** | | | | | | | | | 3. ***Quantitative non-randomised*** | | | | | | | | | | | 4. ***Mixed Methods*** | | | | | | | | |
| --- | --- | --- | --- | --- | --- | --- | --- | --- | --- | --- | --- | --- | --- | --- | --- | --- | --- | --- | --- | --- | --- | --- | --- | --- | --- | --- | --- | --- | --- | --- | --- | --- | --- | --- | --- | --- | --- |
|  | **1.1** | | ***1.2*** | ***1.3*** | | ***1.4*** | ***1.5*** | | ***2.1*** | | ***2.2*** | | ***2.3*** | ***2.4*** | | ***2.5*** | | ***3.1*** | | | ***3.2*** | ***3.3*** | ***3.4*** | ***3.5*** | | | | | ***5.1*** | | | | ***5.2*** | ***5.3*** | | ***5.4*** | ***5.5*** |
| ***Aberdeen*** (1) |  | |  |  | |  |  | |  | |  | |  |  | |  | | ***Yes*** | | | ***Yes*** | ***Yes*** | ***Yes*** | ***Yes*** | | | | |  | | | |  | |  |  |  |
| ***Almutairi*** (2) |  | |  |  | |  |  | | ***Yes*** | | ***Yes*** | | ***Yes*** | ***Yes*** | | ***Yes*** | |  | | |  |  |  |  | | | | |  | | | |  | |  |  |  |
| ***Ballard*** (3) |  | |  |  | |  |  | | ***Yes*** | | ***Yes*** | | ***Yes*** | ***Yes*** | | ***Yes*** | |  | | |  |  |  |  | | | | |  | | | |  | |  |  |  |
| ***Cooper*** (4) |  | |  |  | |  |  | | ***Yes*** | | ***Yes*** | | ***No*** | ***Yes*** | | ***Yes*** | |  | | |  |  |  |  | | | | |  | | | |  | |  |  |  |
| ***Corner*** (5) | ***Yes*** | | ***Yes*** | ***Yes*** | | ***Yes*** | ***Yes*** | |  | |  | |  |  | |  | |  | | |  |  |  |  | | | | |  | | | |  | |  |  |  |
| ***Curyto*** (6) | ***Yes*** | | ***Yes*** | ***Yes*** | | ***Yes*** | ***Yes*** | |  | |  | |  |  | |  | | ***Yes*** | | | ***Yes*** | ***Yes*** | ***Yes*** | ***Yes*** | | | | | ***Yes*** | | | | ***Yes*** | | ***Yes*** | ***Yes*** | ***Yes*** |
| ***Da Silva Serell*** (7) |  | |  |  | |  |  | |  | |  | |  |  | |  | | ***No*** | | | ***Yes*** | ***Yes*** | ***No*** | ***Yes*** | | | | |  | | | |  | |  |  |  |
| ***Degen*** (8) |  | |  |  | |  |  | |  | |  | |  |  | |  | | ***Yes*** | | | ***Yes*** | ***Yes*** | ***Yes*** | ***Yes*** | | | | |  | | | |  | |  |  |  |
| ***Dobbs*** (9) | ***Yes*** | | ***Yes*** | ***Yes*** | | ***Yes*** | ***Yes*** | |  | |  | |  |  | |  | |  | | |  |  |  |  | | | | |  | | | |  | |  |  |  |
| ***Dow*** (10) |  | |  |  | |  |  | | ***Yes*** | | ***Yes*** | | ***Yes*** | ***Yes*** | | ***Yes*** | |  | | |  |  |  |  | | | | |  | | | |  |  | |  |  |
| ***Du Toit*** (11) | ***Yes*** | | ***No*** | ***Yes*** | | ***Yes*** | ***Yes*** | |  | |  | |  |  | |  | | ***Yes*** | | | ***No*** | ***Yes*** | ***No*** | ***Yes*** | | | | | ***Yes*** | | | | ***Yes*** | | ***Yes*** | ***Yes*** | ***No*** |
| ***Ehlman*** (12) |  | |  |  | |  |  | |  | |  | |  |  | |  | | ***Yes*** | | | ***No*** | ***Yes*** | ***No*** | ***Yes*** | | | | |  | | | |  | |  |  |  |
| ***Franzmann*** (13) |  | |  |  | |  |  | |  | |  | |  |  | |  | | ***No*** | | | ***Yes*** | ***Yes*** | ***No*** | ***Yes*** | | | | |  | | | |  | |  |  |  |
| ***Fukuda*** (14) |  | |  |  | |  |  | |  | |  | |  |  | |  | | ***Yes*** | | | ***Yes*** | ***Yes*** | ***Yes*** | ***Yes*** | | | | |  | | | |  | |  |  |  |
| ***Gaugler*** (15) | ***Yes*** | | ***No*** | ***No*** | | ***Yes*** | ***Yes*** | |  | |  | |  |  | |  | | ***No*** | | | ***No*** | ***No*** | ***No*** | ***No*** | | | | | ***Yes*** | | | | ***Yes*** | | ***No*** | ***No*** | ***No*** |
| ***Gerolimatos*** (16) |  | |  |  | |  |  | |  | |  | |  |  | |  | | ***Yes*** | | | ***Yes*** | ***Yes*** | ***Yes*** | ***Yes*** | | | | |  | | | |  | |  |  |  |
| ***Gordon*** (17) |  | |  |  | |  |  | |  | |  | |  |  | |  | | ***Yes*** | | | ***Yes*** | ***Yes*** | ***Yes*** | ***Yes*** | | | | |  | | | |  | |  |  |  |
| ***Guney*** (18) |  | |  |  | |  |  | |  | |  | |  |  | |  | | ***No*** | | | ***Yes*** | ***Yes*** | ***No*** | ***No*** | | | | |  | | | |  | |  |  |  |
| ***Guzman*** (19) |  | |  |  | |  |  | |  | |  | |  |  | |  | | ***Yes*** | | | ***Yes*** | ***No*** | ***Yes*** | ***Yes*** | | | | |  | | | |  | |  |  |  |
| ***Han*** (20) | ***Yes*** | | ***No*** | ***Yes*** | | ***Yes*** | ***Yes*** |  | | |  | |  |  | |  | | ***No*** | | | ***Yes*** | ***No*** | ***No*** | ***No*** | | | | | ***Yes*** | | | | ***Yes*** | | ***No*** | ***Yes*** | ***No*** |
| ***Hanson*** (21) |  | |  |  | |  |  |  | | |  | |  |  | |  | | ***Yes*** | | | ***No*** | ***No*** | ***No*** | ***Yes*** | | | | |  | | | |  | |  |  |  |
| ***Inker*** (22) | ***Yes*** | | ***Yes*** | ***Yes*** | | ***Yes*** | ***Yes*** |  | | |  | |  |  | |  | | ***Yes*** | | | ***Yes*** | ***No*** | ***No*** | ***Yes*** | | | | | ***Yes*** | | | | ***No*** | | ***No*** | ***Yes*** | ***Yes*** |
| ***Isaac*** (23) |  | |  |  | |  |  |  | | |  | |  |  | |  | | ***Yes*** | | | ***Yes*** | ***Yes*** | ***Yes*** | ***Yes*** | | | | |  | | | |  | |  |  |  |
| ***Karacsony*** (24) | ***Yes*** | | ***Yes*** | ***Yes*** | | ***Yes*** | ***Yes*** |  | | |  | |  |  | |  | |  | | |  |  |  |  | | | | |  | | | |  | |  |  |  |
| ***Karel*** (25) |  | |  |  | |  |  |  | | |  | |  |  | |  | | ***Yes*** | | | ***Yes*** | ***Yes*** | ***No*** | ***Yes*** | | | | |  | | | |  | |  |  |  |
| ***Kelleher*** (26) | ***Yes*** | | ***Yes*** | ***Yes*** | | ***Yes*** | ***Yes*** | |  | | |  |  |  | |  | |  | |  | |  |  | |  | | | | |  | | |  | |  |  |  |
| ***Kinderman*** (27) |  | |  |  | |  |  | ***Yes*** | | ***Yes*** | | | ***No*** | ***Yes*** | ***Yes*** | |  | | |  | |  |  |  | | | | |  | | | |  | |  |  |  |
| ***Kirkham*** (28) |  | |  |  | |  |  | | ***Yes*** | | | ***Yes*** | ***Yes*** | ***No*** | | ***Yes*** | |  | | |  |  |  | | | | |  | | | |  |  | |  |  |  |
| ***Livingston*** (29) |  | |  |  | |  |  | | ***Yes*** | | ***Yes*** | | ***Yes*** | ***Yes*** | | ***Yes*** | |  | | |  |  |  | | | | |  | | | |  |  | |  |  |  |
| ***Maidment*** (30,31) | ***Yes*** | | ***Yes*** | ***No*** | | ***Yes*** | ***Yes*** | |  | |  | |  |  | |  | | ***Yes*** | | | ***Yes*** | ***No*** | ***Yes*** | | | | | ***Yes*** | | | | ***Yes*** | ***Yes*** | | ***Yes*** | ***Yes*** | ***Yes*** |
| ***McDermid*** (32) |  | |  |  | |  |  | | ***Yes*** | | | ***Yes*** | ***No*** | ***Yes*** | | ***No*** | |  | | |  |  |  | | | | |  | | | |  |  | |  |  |  |
| ***McGilton*** (33) | ***Yes*** | | ***Yes*** | ***Yes*** | | ***Yes*** | ***Yes*** | |  | |  | |  |  | |  | | ***No*** | | | ***Yes*** | ***Yes*** | ***Yes*** | | | | | ***No*** | | | | ***Yes*** | ***Yes*** | | ***Yes*** | ***Yes*** | ***Yes*** |
| ***Moore*** (34) | ***Yes*** | | ***Yes*** | ***Yes*** | | ***Yes*** | ***Yes*** | |  | |  | |  |  | |  | | ***Yes*** | | | ***Yes*** | ***No*** | ***No*** | | | | | ***Yes*** | | | | ***Yes*** | ***Yes*** | | ***Yes*** | ***Yes*** | ***Yes*** |
| ***Mohr*** (35) |  | |  |  | |  |  | |  | | |  |  |  | |  | | ***Yes*** | | | ***Yes*** | ***Yes*** | ***Yes*** | | | | | ***Yes*** | | | |  |  | |  |  |  |
| ***Mulyani*** (36) |  | |  |  | |  |  | |  | |  | |  |  | |  | | ***No*** | | | ***Yes*** | ***Yes*** | ***No*** | | | | | ***Yes*** | | | |  |  | |  |  |  |
| ***Nakanishi*** (37) |  | |  |  | |  |  | | ***Yes*** | | | ***Yes*** | ***Yes*** | ***Yes*** | | ***Yes*** | |  | | |  |  |  | | | | |  | | | |  |  | |  |  |  |
| ***Noguchi*** (38) |  | |  |  | |  |  | |  | |  | |  |  | |  | | ***No*** | | | ***Yes*** | ***Yes*** | ***No*** | | | | | ***Yes*** | | | |  |  | |  |  |  |
| ***Pieper*** (39) |  | |  |  | |  |  | | ***Yes*** | | | ***Yes*** | ***Yes*** | ***Yes*** | | ***No*** | |  | | |  |  |  | | | | |  | | | |  |  | |  |  |  |
| ***Pinazo-Clapés*** |  | |  |  | |  |  | | ***No*** | | | ***Yes*** | ***Yes*** | ***Yes*** | | ***Yes*** | |  | | |  |  |  | | | | |  | | | |  |  | |  |  |  |
| ***Reinhardt*** (40) |  | |  |  | |  |  | |  | |  | |  |  | |  | | ***Yes*** | | | ***Yes*** | ***Yes*** | ***No*** | | | | | ***Yes*** | | | |  |  | |  |  |  |
| ***Revolta*** (41) | ***Yes*** | | ***Yes*** | ***Yes*** | | ***Yes*** | ***Yes*** | |  | |  | |  |  | |  | | ***Yes*** | | | ***Yes*** | ***Yes*** | ***No*** | | | | | ***Yes*** | | | | ***Yes*** | ***Yes*** | | ***Yes*** | ***Yes*** | ***Yes*** |
| ***Scerri*** (42) |  | |  |  | |  |  | |  | |  | |  |  | |  | | ***Yes*** | | | ***Yes*** | ***No*** | ***Yes*** | | | | | ***Yes*** | | | |  |  | |  |  |  |
| ***Shaw*** (43) |  | |  |  | |  |  | | ***No*** | | | ***No*** | ***Yes*** | ***No*** | | ***Yes*** | |  | | |  |  |  | | | | |  | | | |  |  | |  |  |  |
| ***Sheaff*** (44) | ***Yes*** | | ***Yes*** | ***Yes*** | | ***Yes*** | ***Yes*** | |  | |  | |  |  | |  | | ***No*** | | | ***Yes*** | ***No*** | ***Yes*** | | | | | ***No*** | | | | ***Yes*** | ***Yes*** | | ***Yes*** | ***No*** | ***No*** |
| ***Su*** (45) | |  |  | |  |  |  | ***No*** | | ***No*** | | | ***Yes*** | ***No*** | ***Yes*** | |  | | |  | |  |  | | | |  | | | |  | |  | |  |  |  |
| ***Sung*** (46) | |  |  | |  |  |  | ***No*** | | ***Yes*** | | | ***Yes*** | ***Yes*** | ***Yes*** | |  | | |  | |  |  | | | |  | | | |  | |  | |  |  |  |
| ***Surr*** (47) | |  |  | |  |  |  | ***Yes*** | | ***Yes*** | | | ***Yes*** | ***Yes*** | ***Yes*** | |  | |  | | |  |  | | |  | | | | |  | |  | |  |  |  |
| ***Testad*** (48) | |  |  | |  |  |  | ***No*** | | ***No*** | | | ***No*** | ***Yes*** | ***Yes*** | |  | |  | | |  |  | | |  | | | | |  | |  | |  |  |  |
| ***Torres-Castro*** (49) | |  |  | |  |  |  | ***Yes*** | | | | ***Yes*** | ***Yes*** | ***Yes*** | ***Yes*** | |  | |  | | |  |  | | |  | | | | |  | |  | |  |  |  |
| ***Tropea*** (50) | |  |  | |  |  |  | ***Yes*** | | ***Yes*** | | | ***Yes*** | ***No*** | ***No*** | |  | |  | | |  |  | | |  | | | | |  | |  | |  |  |  |
| ***Williams*** (51) | |  |  | |  |  |  | ***No*** | | | | ***No*** | ***No*** | ***Yes*** | ***No*** | |  | |  | | |  |  | | |  | | | | |  | |  | |  |  |  |
| ***Williams*** (52) | |  |  | |  |  |  | ***Yes*** | | | | ***No*** | ***No*** | ***Yes*** | ***No*** | |  | |  | | |  |  | | |  | | | | |  | |  | |  |  |  |
| ***Yasuda*** | |  |  | |  |  |  |  | |  | | |  |  |  | | ***No*** | | ***Yes*** | | | ***No*** | ***Yes*** | | | ***Yes*** | | | | |  | |  | |  |  |  |
| ***Zhou*** (53) | |  |  | |  |  |  |  | |  | | |  |  |  | | ***Yes*** | | ***Yes*** | | | ***Yes*** | ***Yes*** | | | ***Yes*** | | | | |  | |  | |  |  |  |
| ***Zhou*** (54) | | ***Yes*** | ***Yes*** | | ***Yes*** | ***Yes*** | ***Yes*** |  | |  | | |  |  |  | | ***No*** | | ***Yes*** | | | ***No*** | ***Yes*** | | | ***No*** | | | | | ***Yes*** | | ***Yes*** | | ***Yes*** | ***Yes*** | ***No*** |

**MMAT, 2018 methodological quality criteria:**

**1. Qualitative;** 1.1. Is the qualitative approach appropriate to answer the research question? 1.2. Are the qualitative data collection methods adequate to address the research question? 1.3. Are the findings adequately derived from the data? 1.4. Is the interpretation of results sufficiently substantiated by data? 1.5. Is there coherence between qualitative data sources, collection, analysis and interpretation?

**2. Quantitative RCT;** 2.1. Is randomisation appropriately performed? 2.2. Are the groups comparable at baseline? 2.3. Are there complete outcome data? 2.4. Are outcome assessors blinded to the intervention provided? 2.5 Did the participants adhere to the assigned intervention?

**3. Quantitative non-randomised;** 3.1. Are the participants representative of the target population? 3.2. Are measurements appropriate regarding both the outcome and intervention (or exposure)? 3.3. Are there complete outcome data? 3.4. Are the confounders accounted for in the design and analysis? 3.5. During the study period, is the intervention administered (or exposure occurred) as intended?

**4. Quantitative descriptive;** no studies under this category.

**5. Mixed Methods;** 5.1. Is there an adequate rationale for using a mixed methods design to address the research question? 5.2. Are the different components of the study effectively integrated to answer the research question? 5.3. Are the outputs of the integration of qualitative and quantitative components adequately interpreted? 5.4. Are divergences and inconsistencies between quantitative and qualitative results adequately addressed? 5.5. Do the different components of the study adhere to the quality criteria of each tradition of the methods involved?

**Supplementary Table 3: Characteristics of included Randomised Controlled Trials conducted in care homes, not meeting criteria for higher priority**

| **Study** | **Country** | **Setting/ participants** | **Intervention** | **n** | **Control** | **n** | **Outcome point** | **Outcomes: Kirkpatrick levels** | | | | **Study type** | **VS** |
| --- | --- | --- | --- | --- | --- | --- | --- | --- | --- | --- | --- | --- | --- |
|  |  |  |  |  |  |  |  | 1 | 2 | 3 | 4 |  |  |
| McDermid (32) | UK | Staff in NH with ≥60% residents with dementia | eWHELD; five 25-minute interactive, multimedia online modules &  coaching support fortnightly via video-call/phone from WHELD | 60 Res (29 staff) | eWHELD without coaching | 43 Res (16 staff) | 2 months |  | ☺ |  | ☺ | RCT | 3 |
| Shaw (43) | USA | NH in Kansas, staff & residents | CHAnging Talk (CHAT) staff education program: 3 x 1hr in person, interactive training sessions focused on communication to reduce elderspeak care resistance / BPSD. | 10 NH from RCT with data |  | State data | 6 months |  |  | ☺ |  | 2ndry analysis of RCT data | 2 |
| Williams (52) |  |  |  | 13 NH: 27 staff, 29 Res |  | Cross-over | 1 (PI) & 3 months |  |  | ☺ | ☺ | RCT | 2 |
| Williams (51) |  | 7 NH, all staff | CHAT Online: staff accessed: 3 x 1hr training modules and post-training evaluation online | 4 NH; 141 staff | Waitlist control | 3 NH; 72 staff | PI |  | ☺ | x |  | RCT | 1 |
| da Silva Serelli (7)* | Brazil | 2 CH, staff & PLWD | Portuguese STAR; 12 modules with lectures, discussion, games, video vignettes & handouts, to reduce BPSD in PLWD. 2 x 2hr group workshops, individual weekly training sessions for staff + meetings with managers; facilitated by OT | 25 staff; 46 PLWD | _ | _ | PI (3 months) |  |  |  | ☺ | Quant | 3 |
| Pinazo-Clapes (55) | Spain | 2 NH, all staff & residents | Group training on non-pharmacological management of BPSD and psychotropic drugs, 3x 4 hour weekly sessions & 8hrs of private study, on PCC, behaviour orientated approach, communication and emotional-based approach. | 22 staff; 91 PLWD | TAU | 19 staff; 70 PLWD | 6 months (PI), 12, 18 & 24 months |  |  | ☺ | ☺ | RCT | 3 |
| Corner (5) | UK | East England CH, all staff & residents | PERSONABLE: 1h workshops with reflective exercises, by a mental health nurse/researcher. | 3 CH (44 staff) | TAU | 3 CH (40 staff) | 10 weeks | ☺ |  |  |  | RCT | 3 |
| Testad (48) | Norway | 24 CH, all staff & residents | Trust Before Restraint intervention; Delivered by research nurses: 2 day seminar, then 1 hour monthly groups over 6 months around alternatives to restraint & psychotropic drugs | 118 res | TAU | 156 res | 7 months (PI) |  |  | x |  | RCT | 2 |
| Tropea (50)* | Australia | 24 NH, staff & PLWD | IMPETUS-D; 11 online or face-to-face modules, each 15-45 mins, on palliative and end of life care for PLWD, over 6 months. | 12 NH; 608 PLWD | TAU | 12 NH; 696 PLWD | 6 & 12 months |  |  |  | x | RCT | 3 |

☺: statistically significant outcome; x: no significant outcome; BPSD: Behavioural and Psychological Symptoms of Dementia; CHAT: CHanging Talk intervention; CH: Care Home; IMPETUS-D: Improving Palliative care Education and Training Using Simulation in Dementia; NH: Nursing Home; OT: Occupational Therapist; PERSONABLE: Phase one intervention development: The training intervention; Quant: Quantitative non-randomized study; RCT: Randomized Control Trial; STAR: Staff Training in Assisted Living Residences; VS: Validity Score;

**Supplementary Table 4: Characteristics of included non-RCT studies conducting in care homes (Validity score 4+)**

| **Study** | **Country** | **Setting/ participants** | **Intervention** | **n** | **Control** | **n** | **Outcome (from baseline)** | **Outcomes: Kirkpatrick levels** | | | | **Study type** | **VS** |
| --- | --- | --- | --- | --- | --- | --- | --- | --- | --- | --- | --- | --- | --- |
|  |  |  |  |  |  |  |  | **1** | **2** | **3** | **4** |  |  |
| Zhou 2022 (53) | China | All staff, NH in Jiangsu province (2020-1), with ≥ 10 residents with dementia/ 10 staff | DEmentia Competence  Education for Nursing Home Taskforce (DECENT) Programme: 8 in-person groups from trained educator weekly | 5 NH (101 staff/ Res pairs | Printed material only | 5 NH (116 staff/ Res pairs) | 2 & 3 months |  | ☺ |  |  | Quant | 5 |
| Zhou 2022 (54) |  | All staff, 1 NH in Nanjing |  | 1 NH (12 staff) | _ | _ | 2 months | ☺ | ☺ |  |  | MM | 4 |
| Maidment (30,31) | UK | CH in West Midlands area with ≥40 Res; All staff/ Res with dementia & psychotropic medication | MEDREV: An interactive face-to-face 3-hour educational PCC group (n≤16) workshop (from health psychologist) for care staff; then senior pharmacist psychotropic medication review | 5 CH (142 staff, 34 Res) | - | - | 2,3 (PI), 6 months | ☺ |  |  |  | MM | 5 |
| Guzman (19) |  | 1 NH, 1 CH, all staff & residents | CCSEP; 3 day theatre/ film-based staff training, by coach & two actors, staff develop ideas, make film with Res/ families; reflect on culture change. | 106 staff | - | - | Day 3 (Post-intervention) |  | ☺ |  |  | Quant | 4 |
| Moore (34) |  | 2 NH in London: staff, family carers, people with advanced dementia | Interdisciplinary care leader (clinical supervised) led: (1) integrated, interdisciplinary assessment and care of Res with advanced dementia; (2) education and support for paid and family carers. | 9 PLWD, 4 family carers, 28 staff | - | - | 2,11,15 months | ☺ |  |  |  | MM | 5 |
| Revolta (41) |  | Staff caring for PLWD across health and social care settings | BPS model of dementia involving an interactive group training session (4 hours) and workshop (2.5 hours) facilitated by researcher. | 11 CMHT,  8 wards, 18 CH staff | - | - | 4 weeks (PI) |  | ☺ |  |  | Quant | 5 |
| Fukuda (14) | Japan | Staff from RH | 2 hour educational program focused on guidelines for initial coping with BPSD | 9 RH; 185 staff | TAU | 8 RH; 172 staff | 6 months |  |  |  | ☺ | Quant | 5 |
| McGilton (33) | Canada | Staff from 1 CH | development of individualised communication plans, a 4 hour dementia care workshop and support on the implementation of the communication plan | 20 staff, 12 Res | _ | _ | 10 weeks (PI) |  | x |  | ☺ | MM | 5 |
| Hanson (21) |  | Staff & physicians from 170 CH | Education to reduce antipsychotic medication prescribing | 217 staff; 26 family members | _ | _ | 3 years | ☺ |  | ☺ |  | MM | 5 |
| Dobbs (9) | USA | Certified nursing assistants from 10 NH | Online training, 10 x 1hr modules applying the CARES concepts to ADL for PLWD, focusing on PCC | 48 staff | _ | _ | PI |  | ☺ |  |  | MM | 5 |
| Gerolimatos (16) |  | All staff, 1 CLC | 8 hour, 1 day workshop on dementia using video, lecture and role-plays, primarily online | 105 staff | _ | _ | PI (4 months) |  | ☺ |  |  | Quant | 5 |
| Gordon (17) |  | Staff & MDT, NH | Project ECO-AGE: Bi-weekly, 120min, video conference sessions on managing dementia and behaviour, for 18 months | 11 NH | TAU | 22 NH | 6, 18 months |  |  | ☺ |  | Quant | 5 |
| Reinhardt (40) |  | Direct care workers & MDT in NH | 6 day training on a Person Directed Care (PDC) model, empowering DCW. | 2 NH; 72 PLWD | TAU | 3 NH; 72 PLWD | 6 months |  |  |  | ☺ | Quant | 4 |
| Aberdeen (1) | Australia | All CH staff | Trained team leaders/ facilitators and introduced Team concept mapping over 9 months to enhance multi focal PCC & planning & team learning. | 123 staff; 11 CH | TAU | 25 staff; 4 CH | 9 months |  | ☺ |  |  | Quant | 5 |
| du Toit (11) |  | Nurses and care managers from 9 RH | Peer Enablement Program; Problem solving to promote PCC approach to BPSD; 3 x 2 day workshops over 3 months, Dementia Australia facilitators | 181 staff | _ | _ | PI | ☺ |  |  |  | MM | 4 |
| Isaac (23) |  | 5 Rural NH: staff & PLWD | Harmony in the Bush; 2 hour workshop in 3 weeks with a PCC plan and personalised music playlists used with PLWD for 20-30 mins 2x a day/ 8 weeks | 87 staff; 74 PLWD | _ | _ | PI |  |  | ☺ | ☺ | Quant | 5 |
| Karacsony (24) |  | All staff from 1 RH | 2 x 2hr training in 3 days on knowledge of dementia illness trajectory, pain and symptom management, and Namaste Care Program (focuses on the 'loving' touch and a calm environment) | 35 staff |  |  | PI |  | ☺ |  |  | MM | 5 |
| Degen (8) | Germany | Care workers from NH | MultiTANDEM plus; train the trainer to deliver TANDEM communication training, 6 x 6hr sessions & 3hr session where techniques are evaluated. | 8 NH; 226 PLWD | TAU | 6 NH; 132 PLWD | 21 months |  |  | ☺ | ☺ | Quant | 5 |
| Scerri (42) | Malta | All staff at Public RH & NH | 7x 2hr mainly didactic, large group (approx. n=20) program: dementia management & care | 214 staff | _ | _ | 4 months (PI) |  | ☺ |  |  | Quant | 4 |

NH: Nursing Home; DECENT: Dementia Competence Education for Nursing Home Taskforce; Quant: Quantitative non-randomized study; Res: residents; ☺: statistically significant outcome; x: no significant outcome; MM: Mixed Method Study; CH: Care Home; CCSEP: The Ladder to the Moon Culture Change Studio Engagement Programme, BPS: Biopsychosocial model; PLWD: Persons Living With Dementia; CMHT: Community; Mental Health Team; RH: Residential Home; ADL: Activities of Daily Living; BPSD: Behavioral and Psychological Symptoms of Dementia; CLC: Community Living Centre; CARES: CARES® Dementia-Related Behavior™ Online Training Program (or CARES® Behavior); PCC: Person Centered Care; PI: Follow-up immediately post-intervention; TAU: Treatment as Usual; VS: Validity Score; DCW: Direct Care Worker; MDT: Multidisciplinary team of professionals who support residential facilities.

**Supplementary Table 5: Characteristics of included non-RCT studies conducting in care homes (Validity score <4)**

| **Study** | **Country** | **Setting/ participants** | **Intervention** | **n** | **Control** | **n** | **Outcome point** | **Outcomes: Kirkpatrick levels** | | | | **Study type** | **VS** |
| --- | --- | --- | --- | --- | --- | --- | --- | --- | --- | --- | --- | --- | --- |
|  |  |  |  |  |  |  |  | 1 | 2 | 3 | 4 |  |  |
| Franzmann (13) | Germany | Nurses and care workers from 21 NH | Train the trainer program for senior staff to deliver the TANDEM intervention to foster communication skills in care workers. 2 hr sessions every 2 weeks for 6 months supporting qualified staff to develop, deliver, and evaluate TANDEM training modules | 14 NH; 116 nurses/ care workers | Project staff delivered training to staff directly | 7 NH, 1 hospital ward. | 6 months (PI) |  | x |  | ☺ | Quant | 3 |
| Mulyani (36) | Indonesia | All staff from 1 long term care facility | 2-day, 18hr, by certified trainers. understanding dementia, PCC, communication, BPSD, meaningful engagement. Lectures, discussion, role play | 30 staff | _ | _ | PI |  | ☺ |  |  | Quant | 3 |
| Noguchi (38) | Japan | Staff from 12 CH | 2 x 120 min sessions over 2 days, then care plans implemented by staff members over a month | 35 staff; 14 PLWD | _ | _ | PI: 2 months |  |  |  | ☺ | Quant | 3 |
| Yasuda (56) |  | Care staff from 1 NH | Three sets of 60-90min DCM and dementia training, with 10-15 staff participating at once | 40 staff 40 PLWD | _ | _ | 1 month (PI) |  |  |  | ☺ | Quant | 3 |
| Guney (18) | Turkey | Staff from 2 NH | 8 week program training staff to make life story booklets for PLWD and "About Me" room profiles | 32 staff | _ | _ | PI |  | ☺ |  | x | Quant | 2 |
| Sheaff (44) | UK | Staff from 13 care homes | Dementia Champion trained other staff in dementia awareness and change management through ‘Plan-Do-Study-Act’ cycles | 298 staff | TAU | 10 care homes, 245 staff | Pre-test, PI |  |  |  | ☺ | MM | 3 |
| Han (20) | USA | Staff from residential care settings | Compassionate Touch Program involved 2 hours of coaching on dementia specific CT and communication techniques | 112 staff | _ | _ | PI | ☺ |  |  |  | MM | 3 |
| Ehlman (12) |  | All staff from 12 NH | 7 hr, online, training then 2 day workshop. neurological changes in dementia and focus on retained skills | 22 nursing workers | _ | _ | PI |  | ☺ |  |  | Quant | 3 |
| Gaugler (15) |  | Care workers: NH/ ALF | CARES online training including 4 modules (4hrs) over one month | 40 care workers | _ | _ | PI | ☺ | ☺ |  |  | MM | 2 |
| Hanson (57) |  | All staff from 3 NH | "Comfort First" web-based 6 x 20 min modules, evaluation & implementation guide | 146 staff | _ | _ | PI |  | ☺ |  |  | Quant | 2 |
| Inker (22) |  | Staff and managers from 9 NH | 52 week, 52 lesson online micro-learning curriculum, each lesson averaged 6 mins & 2 question quiz, visual and audio elements | 130 staff | _ | _ | PI | ☺ | ☺ |  |  | MM | 3 |

☺: statistically significant outcome; x: no significant outcome; ALF: Assisted Living Facility; BPSD: Behavioural and Psychological Symptoms of Dementia; CARES: CARES® Dementia-Related Behavior™ Online Training Program (or CARES® Behavior); CH: Care Home; DCM: Dementia Care Mapping; MM: Mixed Method Study; NH: Nursing Home; PLWD: Persons Living With Dementia; PCC: Person Centered Care; PI: Follow-up immediately post-intervention; Quant: Quantitative non-randomized study; Res: Residents; TAU: Treatment as Usual; VS: Validity Score; CT: Compassionate Touch.

***Supplementary Table 6: Comparing Home Care Interventions prepared for stakeholder consultations***

|  | NIDUS-professional | PITCH | EGM- VR (Sung) | BASE (Nakanishi) |
| --- | --- | --- | --- | --- |
| Who was it targeted at? | HCW | HCW | HCW | Care managers & HCW |
| What was it? | 6 manualised sessions, interactive and practice-based, then 3 reunion groups | 2 in-person sessions or 3 online interactive, practice-based sessions, and web resources | 8 x interactive, 15 min e- modules, monthly in-person 1hr supervisor mentoring in groups of 8-10, including VR, social media support; | 2 day training, HCW used web-based tool to identify client unmet needs, interdisciplinary conference for client |
| How was it delivered? | video call groups 6-8 HCW | In person or online groups, in agencies, groups with 10-12 HCW | E-learning  In-person/ VR in groups  Social media | 2-day in person training, recording client behavior in web tool, case meeting for client |
| Who by? | 2 facilitators without formal clinical training | 1 experienced trainer | Home care supervisor | Expert trainers |
| How many hours? | 8 hours | 6 hours (+ travel) | 5 hours (+ travel) | 8 hours initial training then >1 hour per client |
| Over how long? | 6 months | 6 months | 3 months | 6 months |
| Were Tier 2 topics covered? | Yes, manualised | Yes, manualised | Yes, e-modules | Less structured: Behavior management and person-centered care focus |
| Were additional resources needed? (are costs known?) | Clinical psychology supervision; £137/client | Not yet reported |  | In Japanese system, monthly meetings with care manager mandatory |
| What were the results? | Acceptable, perceived as useful (Level 1) | Improved sense of competence, attitudes and knowledge (Level 2) | Improved knowledge, attitudes, competence, empathy (Level 2)* | Reduced challenging behaviors |

Behaviour Analytics & Support Enhancement (BASE); Home Care Worker (HCW)

*when VR included, compared with the intervention without VR; in a lower quality study, the supervision groups and e-modules improved these outcomes relative to no additional training

**Supplementary Table 7: Comparing the interventions found to be effective in care homes**

|  | **What was it?** | **How was it delivered?** | **Tier 2 topics covered?*** |
| --- | --- | --- | --- |
| **WHELD** (3) | In-person, care staff training in PCC, tailored activities, social interactions & system for triggering appropriate antipsychotic medication reviews by the prescribing physician | 9-month program: WHELD therapist (OT) (1 per 9 NH) visited staff and residents for 2 days, identified WHELD champions received 1 day training per month, offsite (months 2-5), onsite (months 6-9) to **cascade** training to staff, implementing flexibly | Yes, not 9-12 |
| **e-WHELD** (32) | Self-directed multimedia e-modules & virtual coaching support for care staff | 2-month program: 5x 25min e-modules, aiming for one a week; fortnightly virtual coaching by researcher |  |
| **STAR-VA**(6,25,35) | National Consultants train local Site-Leads who train staff in expectations, communication, management of BPSD | Site nurse (NC) & behavioural coordinators (BC – psychologist/psychiatrist), attend 3-day workshop then **cascade**; 6 months of group consultation calls (weekly - BCs / 1-2 monthly - NCs) | Yes but not 7,9- 12 |
| **CHAT & CHATO** (43,51,52) | Care staff training focused on communication to reduce elderspeak, care resistance and BPSD. | 3 x 1 hour interactive training modules over 3 weeks: on-site, in person, or online. Multiple sessions to be flexible for staff schedules. | Yes but not 9- 12 |
| **OPAL** (28) | Training for MDT, covering BPSD | 5 month program: 90 minute in-person session for MDT then 3, monthly interdisciplinary video call discussions of 4-5 cases. Delivered by investigators | Yes, not 9- 12 |
| **MMC** (2) | E-learning and video-conferences with staff, medication audits and informational resources on non-pharmacological strategies and reducing antipsychotic use | 6-month program: 1 hour online video Training & 8 key staff video conferences with Dementia Training Australia consultants, every 2-4 weeks. | Yes not 9- 12 |
| **PROCUIDA-Demencia** (49) | Manualized psychosocial intervention for staff (MDT and care home) and managers: PCC, reminiscence, dance & doll therapy and anti-psychotic drug review. | 2-day, on-site group intervention with staff, residents & their relatives at care homes, delivered by study team. Specialist psychologist and psychiatrist for additional training for physicians and nurses. | Yes not 9-12 |
| **STA OP!** (39) | Training for MDT, to enhance physical and affective assessment skills that targeted unmet needs of PLWD. | 5 x 3hr meetings of MDT; training was linked to structured daily/weekly team meetings. The project coordinator visited weekly. | Yes not 9-12 |
| **Pinazo-Clapés** (55) | Care staff training on non-pharmacological management of BPSD and psychotropic drug prescribing protocols | 3 x 4 hour weekly group training for care staff, by  psychologist and an individual trained in PCC; and 8 hours of self-directed learning | Yes |

BPSD: Behavioural and Psychological Symptoms of Dementia; CHAT(O): MDT: Multi-Disciplinary Team who provide health and social care to residents; MMC: Medication Management Consultancy; OPAL: Optimizing Prescribing of Antipsychotics in Long-term care; PCC: Person- Centred Care; PLWD: People Living with Dementia; STAR-VA: Staff Training in Assisted Living Residences (adapted from STAR by Veteran’s Health Administration); WHELD: Well-being and Health for People Living with Dementia

*based on evidence in publications

**Supplementary Table 8: Key policy documents influencing dementia training in Social Care since 2015**

| Date | Author | Report name | Key points relevant to dementia training in social care |
| --- | --- | --- | --- |
| February 2015 | Department of Health (58) | Prime Minister’s challenge on dementia 2020 (59) | Aspirations: (1) Newly appointed social care support workers to have training as part of Care Certificate, with the Care Quality Commission asking for evidence of compliance with the Care Certificate. Expectation that social care providers provide appropriate training to all other staff. (2) Social care staff should be supported to spot early symptoms of dementia and helping them access high quality care and support. (3) Evidence base for what works in dementia training, to develop smarter education and training programme |
| November 2016 | Alzheimer’s Society | Fix Dementia homecare(60) | Recommendations were:   1. Minimum training standards for dementia, framed through the Care certificate, should be raised to align with Tier 2 of the Dementia core skills education and training framework. 2. The Care Quality Commission should include checks for quality of dementia training in their inspection framework for homecare providers. 3. All homecare providers should have an identified, dedicated dementia lead. 4. Every local authority should have a dementia-specific training programme for care workers operating within their borough. 5. Government bodies must find the required resource to enable all homecare workers to receive dementia training. |
| 2015, updated 2018 | Skills for Health, Health Education England and Skills for Care | Dementia Core Skills Education and Training Framework(61) | Benchmarks dementia training for the health and care workforce to support staff to have the right knowledge and skills to deliver high quality, person- centered care for people living with dementia and their carers. It comprises three Tiers, including awareness, basic skills and leadership. These reflect the role and degree of contact different staff have with people living with dementia.It suggests Tier 1 dementia awareness training in first 3 months of appointment.staff working regularly with people living with dementia and expert leaders undertake Tier 2 or 3. |
| June 2018 | National Institute for Health and Care Excellence (NICE) | Dementia: assessment, management and support for people living with dementia and their carers  NICE guideline 97 | Section 1.13 (Staff training and education) states all staff should have training in person centred and outcome-focused dementia care; with additional face-to-face training and mentoring to staff who deliver care and support to people with dementia, including training on understanding, reacting to and helping people who experience behaviours indicating distress, with follow-up sessions to discuss specific situations, advice on interventions that reduce the need for antipsychotics, and promote freedom of movement. Training should include starting and holding difficult and emotionally challenging conversations. |
| June 2019 | National Institute for Health and Care Excellence (NICE) | Dementia  Quality standard [QS184] | Quality statement 7, Carers of people with dementia are offered education and skills training; applies to community care providers |
| June 2019 | All-Party Parliamentary Group (APPG) on Dementia | Hidden no more: dementia and disability (62) | Report about dementia awareness and skills across the broad social workforce. Reported concerns of inadequate training. Recommendation that all home and residential care staff should receive mandatory training. This should be equivalent to Tier 2 of Dementia Core Skills Education and Training Framework, provided by Health Education England. |
| June 2021 | NHS England | Dementia wellbeing in the COVID-19 pandemic | Outlined staff training and support needs related to the pandemic, including in the use of remote techniques. |
| December 2021 | Department of Health and Social Care (DHSC) | People at the Heart of Care: adult social care reform white paper (63) | Announced planned investment of £500 million for a knowledge and skills framework, careers pathways and learning and development to support progression for care workers and registered managers, fund Care Certificates, with improved portability when moving roles; £50m per year was committed to fund the Care Certificate |
| February 2022 | Department of Health and Social Care (DHSC) | Joining up care for people, places and populations: health and social care integration white paper (64) | Sets out the government’s ambition to accelerate the delivery of joined-up health and social care, reduce barriers to staff moving between the NHS and social care, opportunities for cross-sector training and joint roles, increase the number of learning experiences in social care and consider the introduction of an integrated skills passport |
| February 2022 | NHS England | The Well Pathway For Dementia (65) | The NHS England transformation framework includes “training well”: to develop training that works in social care settings (care homes) as well as health care settings |
| December 2022 | Care Quality Commission (CQC) | Our position on the Care certificate (66) | CQC continues to expect providers to induct, support and train their staff appropriately. Those who employ health care support workers and adult social care workers should be able to demonstrate that staff have, or are working towards, the skills set out in the Care Certificate. This is the benchmark for staff induction. When we start to use our new assessment framework, this will continue to be a key area  Associated guides set out inspection requirements; for care homes, this includes training in communication with people with a learning disability and autistic people, but dementia not specifically mentioned |
| 2023 | Skills for Care | The state of the adult  social care sector  and workforce in England (67) | This report found that 65% of social care staff engaged with the Care Certificate and engagement was highest in domiciliary care services (74% of care workers had engaged with the Care Certificate) in 2022/23; though non-mandatory, 45% had engaged with dementia training. |
| April 2023 | Department of Health and Social Care (DHSC) | Next steps to put People at the Heart of Care (68) | To support implementation of the care workforce pathway – including funding for  a new Care Certificate Level 2 qualification - a baseline standard for all new care workers to work towards |
| June 2023 | NHS England | NHS Long Term Workforce Plan (69) | This will include integrated workforce planning to best develop  and deploy staff; for example, through opportunities for joint teams, joint training and rotation between NHS and social care settings. |
| June 2023 | NHS England | Intercultural Dementia Care Guide (70) | Guidance for health and social care workers on delivering culturally appropriate care, signposted to training resources |
| September 2023 | Department of Health and Social Care (DHSC) | Digitisation is the power to improve care (71) | Outlines plans for a digital care record, and associated training needs |
| August 2023 | Department of Health and Social Care (DHSC) | Major Conditions Strategy (72) | Discusses integrated care, effective prevention releasing significant sums for other services. |
| June 2022 | Department of Health and Social Care (DHSC) | Health and social care review: leadership for a collaborative and inclusive future (73) | Messenger independent report: discussing importance of leadership training and career support in social care |
| May 2023 | Homecare association | Rising demand and hospital discharge - how big are the challenges facing health and social care? (74) | Links social care pressures to delayed discharges |
| November 2022 | ADASS (Directors of Adult Social Service) | There is neither ‘the funding nor the workforce’ needed to meet the needs of older and disabled people this winter | A blog describing economic pressures on the sector |
| November 2019 | Unison College | Skills for the Future(75) | A survey of public sector workers views on training. Social care workers put higher value on training than other sectors |
| October 2023 | Laing Buisson, delivered on behalf of the Care Provider Alliance | Fair cost of Care homes for older people and dementia (76) | Describes North/South inequalities in care, and inequalities between homes with greater and fewer self-funded clients, with care homes rated low by CQC receiving least funding |
| November 2023 | Care England | Bridging The Gap: Tackling Inequalities in England’s Social Care Sector and Beyond | Describes inequalities in the social care sector |
| January 2024 | Department of Health and Social Care (DHSC) | Care workforce pathway for adult social care (77) | new career structure for the adult social care workforce. This is the first phase of the pathway, focusing on direct care roles. It sets our values, knowledge and skills for those new to care, are or support workers, supervisors or team leaders and practice leaders; for dementia it maps to the Dementia Training Standards Framework. |
| [website accessed April 2024] | Skills for Care, Health Education England and Skills for Health. | The care certificate (78) | Agreed set of standards that define the knowledge, skills and behaviours expected of specific job roles in the health and social care sectors. which include awareness of mental health, dementia and learning disability  The Care Certificate is made up of the 15 minimum standards that should be covered if you are 'new to care' and should form part of a robust induction programme. |
| April 2024 | Department of Health and Social Care (DHSC) | Adult Social Care Training and Development Fund: a guide for employers (79) | Launched new fund, including £50 million of the total fund value is dedicated to supporting up to 37,000 individuals in direct care roles to enrol onto the new Level 2 Adult Social Care Certificate qualification by March 2025 |

59.	Cabinet Office. Prime Minister’s challenge on dementia 2020 [Internet]. 10 Downing Street, London: Department of Health and Social Care; 2015. Available from: www.gov.uk/government/publications/prime- ministers-challenge-on-dementia-2020/prime- ministers-challenge-on-dementia-2020

60.	Alzheimer’s Society. Fix Dementia Care: Homecare [Internet]. 2016. Available from: www.alzheimers.org. uk/sites/default/files/migrate/downloads/fix_dementia_care_homecare_report.pdf

61.	Skills for Health, Health Education England and Skills for Care. Dementia Core Skills Education and Training Framework. 2018.

67.	The state of the adult social care sector and workforce in England. Skills for Care; 2021.

76.	LaingBuisson, delivered on behalf of the Care Provider Alliance. Fair cost of Care homes for older people and dementia. 2023 Sep.

77.	GOV.UK [Internet]. [cited 2024 Jul 20]. Care workforce pathway for adult social care. Available from: https://www.gov.uk/government/publications/care-workforce-pathway-for-adult-social-care

78.	Skills for Care. Care Certificate [Internet]. [cited 2024 Jul 21]. Available from: https://www.skillsforcare.org.uk/Developing-your-workforce/Care-Certificate/Care-Certificate.aspx

79.	GOV.UK [Internet]. [cited 2024 Jul 21]. Adult Social Care Training and Development Fund: a guide for employers. Available from: https://www.gov.uk/government/publications/adult-social-care-training-and-development-fund/adult-social-care-training-and-development-fund-a-guide-for-employers
